# “I can’t cope with multiple inputs”: Qualitative study of the lived experience of ‘brain fog’ after Covid-19

**DOI:** 10.1101/2021.08.07.21261740

**Authors:** Caitriona Callan, Emma Ladds, Laiba Husain, Kyle Pattinson, Trisha Greenhalgh

**Author notes:** Corresponding author: E Ladds, Nuffield Department of Primary Care Health Sciences, University of Oxford, Oxford OX2 6GG, UK.

## Abstract

**Objective:** To explore the lived experience of ‘brain fog’—the wide variety of neurocognitive symptoms that can follow Covid-19.

**Design and setting:** UK-wide longitudinal qualitative study comprising online interviews and focus groups with email follow-up.

**Method:** 50 participants were recruited from a previous qualitative study of the lived experience of long Covid (n = 23) and online support groups for people with persistent neurological problems following Covid-19 (n = 27). In remotely-held focus groups, participants were invited to describe their cognitive symptoms and comment on others’ accounts. Individuals were followed up by email 4-6 months later. Data were audiotaped, transcribed, anonymised and coded in NVIVO. They were analysed by an interdisciplinary team with expertise in general practice, clinical neuroscience, the sociology of chronic illness and service delivery, and checked by three people with lived experience of brain fog.

**Results:** 84% of participants were female and 60% were White British ethnicity. Most had never been hospitalised for Covid-19. Qualitative analysis revealed the following themes: mixed views on the appropriateness of the term ‘brain fog’; rich descriptions of the experience of neurocognitive impairments (especially executive function, attention, memory and language), accounts of how the illness fluctuated—and in some but not all cases, resolved—over time; the profound psychosocial impact of the condition on relationships, personal and professional identity; self-perceptions of guilt, shame and stigma; strategies used for self-management; challenges accessing and navigating the healthcare system; and participants’ search for physical mechanisms to explain their symptoms.

**Conclusion:** These qualitative findings complement research into the epidemiology and underlying pathophysiological mechanisms for neurological symptoms after Covid-19. Services for such patients should include: an ongoing therapeutic relationship with a clinician who engages with the illness in its personal, social and occupational context as well as specialist services that are accessible, easily navigable, comprehensive, and interdisciplinary.

**Summary:** *Strengths and Limitations of Study:*

- To our knowledge, this is the largest and most in-depth qualitative study of the lived experience of brain fog in survivors of Covid-19.
- The research team was interdisciplinary and interprofessional, and included consultation with patient experts by experience, who helped with data interpretation and peer review.
- Oversampling from men and non-white ethnic groups allowed partial correction of an initially skewed sample.
- The sample was drawn entirely from the UK
- Residual skews in the samples, particularly regarding minority ethnic groups and occupational classes, limited our ability to capture the full range of experiences

*Funding statement:* This research is funded from the following sources: National Institute for Health Research (BRC-1215-20008), ESRC (ES/V010069/1), and Wellcome Trust (WT104830MA). Funders had no say in the planning and execution of the study or writing up of the paper. KTSP is supported by the National Institute for Health Research Biomedical Research Centre based at Oxford University Hospitals NHS Foundation Trust and the University of Oxford.

*Competing Interests Statement:* EL and TG provided evidence on long Covid for House of Lords Select Committee TG was on the oversight group for the long Covid guideline at the National Institute for Health and Clinical Excellence, and at the time of writing is on the UK’s National Long Covid Task Force. KP and CC have no competing interests to declare.

## Background

It is now well-established that COVID-19 can cause persistent ill-health beyond the acute infection, with results from a representative sample of the UK population suggesting that approximately 1 in 5 people will still experience symptoms 5 weeks after infection, and almost 1 in 7 after 12 weeks [1]. Just under 1 in 10 individuals are still affected after 1 year [2]. Over half of those with ongoing symptoms -termed ‘long Covid’ by patients [3] -experience at least some reduction in ability to carry out their everyday activities, and many report being unable to return to work weeks after the initial infection [1, 4]. The growing number of people with chronic and sometimes disabling illness resulting from the COVID-19 pandemic has made it a policy priority to develop services to meet their health needs [5, 6] and associated clinical and occupational guidelines [7].

Long Covid, a “patient-made” term [3] embraces the formally-defined conditions of post-acute Covid-19 syndrome (symptoms persisting between 4 and 12 weeks) and chronic Covid-19 (symptoms beyond 12 weeks) [7]. It is highly heterogenous in nature, with sufferers reporting a wide range of often-fluctuating symptoms amongst which fatigue, breathlessness, chest pain, post-exertional malaise, autonomic nervous system disruption, and cognitive dysfunction [6, 8, 9] are some of the most common. Whilst the underlying pathophysiology remains unclear, persistent viraemia [10], relapse or reinfection [11] inflammatory and immune reactions [12, 13], deconditioning [14] and psychological factors [15, 16] have all been proposed as contributors. It is likely that in many patients the causative pathways are multifactorial [17].

Analysis of the health records of almost a quarter of a million Covid-19 survivors revealed that neurological and psychiatric presentations occurred in both hospitalised and non-hospitalised patients, affecting around one-third of patients over the following 6 months with most severely affected people at highest risk [18]. Around one-quarter experienced disturbed mood, especially anxiety, and a small fraction developed more serious problems such as psychosis. Other neurocognitive problems included substance use disorder, insomnia, cerebrovascular events, encephalitis, dementia, and disorders of peripheral nerves, nerve roots or plexus [18]. Surveys and focus groups conducted on online samples of mostly non-hospitalised long Covid patients have identified impairments in attentional processing, short-term memory and executive function, alongside a general, befuddled state termed ‘brain fog’ [4, 6, 8, 19]. More recently concern has been raised that such effects may also extend to adolescents and children – a group generally considered to be at ‘low risk’ from Covid-19 infection [20]. A range of possible pathophysiologies have been identified, including direct neuroinvasion [21], viral persistence and chronic inflammation [22], neuronal injury or toxicity and glial activation [21], microvascular injury [23], activation of autoimmune mechanisms [24], and Lewy body production [25] amongst others, with imaging studies demonstrating a differential loss of grey matter in Covid patients in a number of key brain regions [26].

The functional impact of such cognitive impairment is often profound, affecting individuals’ abilities to work and carry out normal daily activities, impeding decision making and judgement, and impairing communication and social relationships, though these impacts have rarely been systematically studied. Guidance for those with neuropsychiatric long Covid symptoms suggests that specialists in clinical psychology and psychiatry should be part of the core multidisciplinary team involved in long Covid rehabilitation [7], but these recommendations are contested and inconsistently implemented. Developments in treatment approaches, service pathways, and occupational support structures require further knowledge of both the mechanistic aetiologies underlying such symptoms as well as a better understanding of the lived experiences of those who suffer them.

In this study, we sought to answer three key questions: a) what neurocognitive symptoms are experienced by adults with long Covid?; b) how do these symptoms impact on individuals?; and c) how do they deal with them? We also sought to explore whether our current understanding of psychocognitive processes and the pathological effects of the Covid-19 virus could inform potential mechanistic explanations.

## Methods

### Study design and governance

This study of people with 50 brain fog was an extension of a previous qualitative study on a large sample of 114 people with long COVID using interviews and focus groups, reported previously [6, 27]. Ethical approval was granted from the East Midlands – Leicester Central Research Ethics Committee (IRAS Project ID: 283196; REC ref 20/EM0128) on 4^th^ May 2020 and subsequent amendments. Participants for the original study had been recruited between May and September 2020 from long Covid support groups on Facebook, a social media call (Twitter), and snowballing (where participants were invited to recruit others known to them). To correct skew, we had oversampled from men and minority ethnic groups. In October 2020, prompted partly by participants’ own desire to explore brain fog further, we emailed everyone in this original sample of 114 asking for volunteers to join additional focus groups, and 23 agreed. To extend the sample, 27 additional participants were recruited from an online support group dedicated to the neurological effects of long Covid. The dataset for the brain fog study thus consisted of selected data from the original interviews with 23 participants plus five new focus groups with the full sample of 50.

Five focus groups were held in October and November 2020; numbers of participants in each group ranged from 10 to 14. Each group had two facilitators who shared the roles of administering and facilitating the group and taking contemporaneous notes. After a brief explanation and affirmation of understanding and consent, participants were invited to tell the story of their neurocognitive symptoms, with conversational prompts to maintain the narrative and elicit information about the impact on an individual’s life and any interaction between symptoms [28]. We encouraged the sharing of stories because the story form is particularly useful for identifying issues important to the patient, identifying emotional touch points in an illness journey, and promoting interaction between participants [29]. One person’s story may attract another similar or contrasting story, and reactions to a story (laughter, anger, sarcasm) can add to the dataset.

### Data management and analysis

Focus groups were audiotaped with consent, transcribed in full, de-identified and entered onto NVIVO software version 12; contemporaneous notes were also entered. Additional material from the original dataset (where people had raised relevant issues) were also included.

In an initial familiarisation phase, sections of text were arranged into nine broad categories:

1. Naming the phenomenon
2. Neurocognitive symptoms
3. Natural history of neurocognitive symptoms in long Covid
4. Fatiguability, and interplay between neurocognitive and physical symptoms
5. Psychosocial impact of persistent neurocognitive symptoms
6. Guilt, shame and stigma related to Long Covid
7. Self-management
8. Navigating the healthcare system
9. Hypothesising mechanisms

An interim synthesis was produced from early transcripts and progressively refined using the constant comparative method (data from each new transcript were used to add nuance to the existing synthesis) [30]. Finally, to add more descriptive depth, clarify any discrepancies or ambiguities within the existing data and to track progression (and perhaps resolution) of symptoms, we sent each participant a follow-up email between 4 and 6 months after the focus group (i.e. 10-12 months after their original Covid-19 illness). We asked how their long COVID symptoms were progressing generally as well as asking them to describe their neurocognitive symptoms in detail. 20 of the participants responded to this email and this data was integrated into, and helped refine, our final interpretation.

### Theoretical framework

Our analysis was informed by three main theoretical lenses.

- First, from a neuroscience perspective in which SARS-COV-2 (the virus responsible for COVID-19) disrupts function in brain and brainstem networks [31] responsible for maintaining body equilibrium (allostasis), adjusting physiological systems (homeostasis) and sensing internal bodily signals (interoception). These systems interact closely with brain systems subserving mood, attention (i.e. fatigue) and cognition [32].
- Second, sociological theories of chronic illness, including May’s burden of illness theory, which focuses on the (sometimes extensive) work needed by patients to manage their illness and navigate the system [33], biographical perspectives on chronic illness, which emphasise the impact of the illness on identity and the role of storytelling in shaping and rebuilding that identity [34-36]; and stigma (the depiction by both self and others of illness as shameful and—at least to some extent—the fault of the person) [37].
- Third, emotional touchpoints of powerful feelings such as anger, fear, or hope [38] were identified in participants’ experiences of healthcare, and experiences engendering strong positive or negative emotions interpreted using theories of good professional practice, including the physician as wise counsel [39], the therapeutic relationship [40] and continuity of care [41].

### Patient involvement statement

The study was planned, undertaken, analysed and written in collaboration with people with long Covid. We gave a webinar presentation via teleconference to which all 50 patient participants were invited, where we presented the key findings including the quotes used in this paper. A recording and copy of the presentation was shared with all participants and all were invited to correct any errors or misinterpretations. The draft paper was modified in response to their feedback. In addition, two clinically qualified people with long Covid reviewed a near-final draft of this paper.

## Results

### Description of dataset

Details of participants are shown in Table 1. Despite our efforts to balance for gender and ethnicity, the final sample was skewed to 42 of 50 (84%) female and 36 (72%) White. By comparison, long Covid support groups are up to 86% female [4] and the UK population is 80-85% White British (depending on how defined) [42]. The 5 focus groups, chat transcripts, and follow-up email communications produced over 1000 pages of transcripts and notes. The nine emergent coding themes are discussed in more detail below with illustrative quotes in Table 2 and definitions of neurocognitive processes/functions in box 1.

**Table 1:** Participant characteristics.

|  | Participants recruited from previous long Covid study | Participants recruited from neuro Covid support groups | Total Brain Fog Focus Group Participants | Responders to email follow-up post-focus groups |
| --- | --- | --- | --- | --- |
|  | 23 | 27 | 50 | 20 |
| Gender<br>Female<br>Male | 15<br>8 | 26<br>1 | 42<br>8 | 17<br>3 |
| Age<br>Median<br>Range | 48<br>31-74 | 36<br>29-68 | 43<br>29-74 | 43<br>31-74 |
| Ethnicity<br>White British<br>White other<br>Black<br>Asian<br>Mixed<br>Non-response | 16<br>3<br>1<br>3<br>0<br>0 | 14<br>3<br>1<br>2<br>0<br>7 | 30<br>6<br>2<br>5<br>0<br>7 | 11<br>1<br>0<br>1<br>0<br>7 |
| Occupation<br>Healthcare professional<br>Non-healthcare professional<br>Non-response | 8<br>13<br>2 | 8<br>11<br>8 | 16<br>24<br>10 | 5<br>9<br>6 |
| Hospitalised at any point due to Covid-19<br>Yes<br>No<br>Non-response | 0<br>9<br>14 | 4<br>8<br>15 | 4<br>17<br>29 | 4<br>16 |

**Table 2:** Participant Quotes.

| Identifier | Source | Quote |
| --- | --- | --- |
| 1 | Participant 10, Focus Group (FG) 4 | “Does anyone ever refer to it as neurocognitive fatigue? In a way I don’t like brain fog as it’s too vague, too loose of a term, so want something more technical. Though I don’t think neurocognitive fatigue encompass the word finding difficulties, so it’s not ideal either” |
| 2 | Participant 7, FG1 | “One of the things I’ve realised is how many things I do in my normal day - I’m not talking about work, just in a normal day - that are cognitive that I [didn’t previously] think of as being cognitive. So a supermarket, the amount of sensory information, and just staring at a row of things looking for the food that you want, remembering where things are in the aisles and planning your trip so that you don’t have to walk backwards and forwards around the shop, that surprised me. [...] Not just can I walk around the supermarket, it’s planning, it’s getting there, it’s choosing stuff, all of that is actually really difficult.” |
| 3 | Participant 5, FG1 | “I can’t cope with multiple inputs, like if I’m trying to reply to a message on my phone and one of my boys starts speaking to me or there’s something else happening as well that just really fries my brain. I mean I used to be the kind of person that, like all |
|  |  | women, multi-tasking was a superpower. I was able to, do lots and lots of things, you know I'm [a doctor]; I would have one patient I'd be hearing lots about another patient coming I'd be remembering I'd be doing something else I'd be juggling lots and lots of things and now I can't keep multiple plates spinning I absolutely can't. I've got to focus on just one thing or I make massive mistakes and it's like I forget my intentions all the time." |
| 4 | Participant 10, FG3 | "I can ask somebody a question and then I'll ask the exact same question two minutes after and not remember I've asked them, I can't remember significant things that have happened in the past either" |
| 5 | Participant 8, FG2 | "[It's difficult] to comprehend and take in written information and read it. I had a form sent to me at work and I just felt, 'I can't do this at the moment' and put it to one side and hoped to come back to it because it's just been too difficult" |
| 6 | Participant 3, FG5, in email response to follow-up | "I'm probably about 90% better. I'm struggling to put in full days at work and still need a great deal of rest and sleep. My brain fog is greatly improved, although I'm making mistakes at work and have been forgetful and sometimes confused with large amounts of new information. I feel like my head is clear now. When you did the group interview I felt like I was drugged up all of the time. Now it's far and few days between that I feel that way. I think the brain fog lasted around eight months." |
| 7 | Participant 9, FG1 | "I've had times when I've tried to do teaching or have meetings via Zoom or just spent a lot of time doing computer work, then I'd |
|  |  | often relapse the following day, what I get is burning lungs, chest pain, breathlessness and the tachycardia. So without a doubt the mental exertion or the energy required to do the thinking and processing then has a detrimental effect on me physically” |
| 8 | Participant 11, FG3 | “Seven months plus in I don’t know whether I’m gonna get my brain back [...] I’m really, really fearful for the future or whether I’m going to be able to get back to what I want to do and that’s like your identity and yourself and who I am as a person is, you know, a big part of me is being a [allied health professional] and if I can’t, if I’ve lost that, I’ve lost a huge part of me.” |
| 9 | Participant 9, FG4 | “I found myself restating and reiterating many times professionally where I’m at now in terms of cognitive ability and there’s only so many times you can do that before I feel like I’m becoming that person, you know and it’s a lot easier to do that in the house but I think professionally it’s been really hard” |
| 10 | Participant 5, FG4 | “a few times that I’ve been out and had an in-depth conversation with somebody that hasn’t managed to get used to how I am, they’ve sort of said to me “you’re going round in circles in your conversation” or “you’re not making a lot of sense”, when I hadn’t quite recognised how repetitive I was being until somebody said it back to me. But even so those same people ... can’t seem to cut me any slack for it, or can’t seem to understand how difficult it is, do you know what I mean? [There] just doesn’t seem to be the understanding there and I can understand that because it would be beyond my comprehension as well if I hadn’t lived it” |
| 11 | Participant 5, FG2 | “For me it’s been going from working at 110% pace to not being able to get out of bed, not being able to work to not see people, to have to cancel plans, the impact on my life has been a massive transition and getting my head around that has been huge. I’m accepting now that I need to take the time off to get better and although that’s really difficult and it’s meant letting lots of people down, and there’s been a complete change in my life, I’ve managed to get to that place.” |
| 12 | Participant 7, FG4 | “Me and my husband have got a traffic light system now, so green’s fine, he can just talk business at me, amber is like can you just keep ‘what’s the weather’-like kind of conversation, and then red is just stop, I need to just rest, stop all the sensory input coming in. And that seems to be working quite well now, so literally I’ve got to say amber or red and it’s that thing when you’re so tired that you can’t even articulate that you’re so tired and explain. So that really has helped us and I think might stop quite a lot of rows.” |
| 13 | Participant 2, FG1 | “I’ve gone to the GP’s and it’s like I speak to someone different every time which is not helpful for that continuity and that consistency and it’s like I have to go right back to the beginning and it’s almost like I’m not believed that I even had Covid, and it’s like I’m so far away from my normal and it’s that trickiness of having to re-explain in that ten minutes and then you just go bluurrghh and it like all comes racing out and you’re like ‘I’ve |
|  |  | no idea what I've just said, a) whether it makes sense or b) whether I actually got my point across''' |
| 14 | Participant 8, FG1 | "I have to say it was when my GP said 'yes, we recognise what you've got as Long Covid and we're treating it like concussion at the moment until we know more about it, and we will recommend you rest and maybe try these drugs', I mean, I almost broke down it was the acknowledgement of the issue. [It] takes away so much of the stress because, we're all [thinking], you know, 'is this really happening, is this just me malingering or do I really have this thing'. And so that was that was a key moment for me" |
| 15 | Participant 7, FG 1 | "I had a couple of different GPs that I spoke to at the beginning and then I spoke consistently to the same locum GP and she was very good. It was when I was having quite a difficult time trying to go back to work and I was struggling quite a lot psychologically and she was very supportive, she spent a lot of time with me and that consistency was good" |
| 16 | Participant 13, FG2 | "I've treated stroke patients who [have] dysphasia and they can't find the right words so they go around the houses to describe something so that you understand what they mean and it felt a bit like that in a way that you know what you want to say but you can't think what that word is because it doesn't come to the forefront of your mind. So you're trying to think of how you can describe it and I thought 'oh gosh, I've turned into one of my |
|  |  | stroke patients' because I'm trying to find another suitable word<br>but it's such a struggle though" |

### Naming the phenomenon

Participants varied in their attitudes towards the patient-made term ‘brain fog’ [4]. Some found it useful as an accessible shorthand to disclose their wide-ranging cognitive difficulties to others, but others felt the term lacked specificity or did not adequately convey the severity of their symptoms (Quote 1).

### Neurocognitive symptoms

Participants’ description of the symptoms and functional impairments of brain fog were often consistent with deficits in specific domains of cognitive function—particularly executive function, attention, memory and language. Deficits in executive function included problems with planning, decision-making, flexibility and working memory (Quote 2), whilst impairments in complex attention included difficulties with selective, sustained attention, divided attention, and processing speed (Quote 3), and long-term memory impairments were seen in free recall, cued recall, procedural memory, and autobiographical memory (Quote 4). The specific language deficits experienced by focus group participants varied between individuals, including difficulties with word-finding and fluency, syntax, reading comprehension and writing (Quote 5).

### Natural history of neurocognitive symptoms in long Covid

The longitudinal nature of the study allowed us to explore some aspects of progression of the condition. In the email follow-up, a majority of participants reported that their brain fog symptoms had only become evident after their initial acute Covid illness, with the delay of onset ranging from one to four months, and a majority of participants having ongoing but improving brain fog symptoms at time of follow-up. Of those who felt their brain fog had resolved entirely, the range of time to resolution of symptoms after initial acute illness was 6-10 months (note, however, that this study was not designed to identify precise time course). In those whose symptoms of brain fog persisted, however, these tended to fluctuate both throughout the day and also over a timescale of weeks to months, typically, but not invariably, showing gradual long-term improvement (Quote 6).

### Fatiguability, and interplay between neurocognitive and physical symptoms

Fatiguability featured prominently, with many participants describing how either physical or mental effort precipitated a decline in their neurocognitive symptoms. There was also clear interplay between physical and cognitive symptoms, with physical fatigue, tachycardia, or breathlessness most frequently described as impacting the latter (Quote 7).

### Psychosocial impact of neurocognitive symptoms

Participants described profound psychological and social impacts, notably inability to return to work at their previous functional level or even at all. Participants who had returned to work described how they now had reduced hours or adapted roles (e.g. relying on others to check their work), which were often associated with anxiety about potential risks associated with their mistakes (Quote 3), self-doubt about their own abilities, loss of self-worth and altered identity (both professional and personal), as illustrated by Quote 8.

### Guilt, shame and stigma

Participants frequently reported strong emotional responses induced by their symptoms and in others’ reactions to them. Guilt and shame were particularly evident and often related to difficulties returning to work or their previous level of function or a lack of understanding from others about these problems (Quotes 9 & 10). Particularly troubling were deficits that were not physically visible to other people, and which in some contexts they felt they had to conceal, such as difficulties with language or memory. Participants also described instances of interpersonal conflict arising from their varying cognitive function (Quote 12).

### Self-management

Many participants had developed coping strategies to deal with their symptoms, principally around lowering self-expectations and prioritising rest. This resulted in complex self-negotiations and activity trade-offs, including limiting return to work, which participants found frustrating and psychologically draining (Quote 11). Moreover, communication of their reduced, and often varying, cognitive function to family, friends or colleagues, was a significant challenge, thus some participants had developed innovative strategies to try and convey their current symptoms and level of functioning (Quote 12)

### Navigating the healthcare system

Participants had varying experiences of navigating systems of healthcare, with neurocognitive symptoms often adding to the difficulties of communicating and self-advocating with healthcare professionals (Quote 13). Many described strong negative emotions of frustration, anger and hopelessness associated when they perceived healthcare professionals as having dismissed their symptoms as ‘in your head’, secondary to depression or anxiety, or not real. Conversely, some participants described a sense of huge relief and validation at feeling believed and having their symptoms acknowledged -often framed as a small victory in the overall uncertainty of long Covid (Quote 14). This was particularly true in the context of interactions with healthcare practitioners, where continuity, wise counselling, and bearing witness were also heralded as desirable components of effective therapeutic relationships (Quote 15).

### Hypothesising mechanisms

Participants frequently attempted to make sense of their symptoms and communicate the severity and legitimacy of their suffering through analogous referral to disorders such as stroke, concussion or dementia (Quotes 14 & 16). Many had undergone investigations without identifying a clear cause; in such cases in particular, participants were keen to hypothesise about the physical or neuropsychiatric mechanisms for their as yet unexplained symptoms. Some reported trialling various strategies of self-management, sometimes based on hypothetical mechanisms of long Covid they had read about. These included: dietary adaptations – eg: low histamine trials, food supplements eg: zinc, or complementary therapies eg: cannabinoid oils, and were met with varying success.

## Discussion

### Summary of key findings

This qualitative study of 50 people in the UK suffering from neurocognitive symptoms (brain fog) following Covid-19 has revealed several important findings. Common symptoms in this sample included deficits in executive function, attention, memory and language, which may not be seen – or noticed – until several weeks to months after the acute viral illness, and in most cases followed relapsing-remitting course generally with gradual improvement over several months. Prominent fatiguability and interaction between cognitive and physical symptoms combined with the psychosocial impact on professional and personal activities to produce a destabilising, debilitating, frustrating, stigmatising and frightening situation, impairing individuals’ functional ability and damaging their personal and professional identity. They used various approaches to mitigate the effects of brain fog including activity trade-offs and communication strategies, but despite this had only limited success. The experience of illness was greatly compounded by the challenges experienced in navigating the healthcare system—a task which required the very neurocognitive skills they currently lacked.

### Comparison with theoretical literature

Some accounts of the varied, uncertain and non-linear nature of this condition fitted Frank’s definition of the ‘chaos narrative’, where the illness experience is unresolved by restitution of the former healthy self, thus remains confusing and lacking in meaning [36]. The profound impact of symptoms on individuals’ independence, self-efficacy, and self-trust resonated with previous descriptions of spoiled identity and the disrupted sense of purpose and self that can accompany chronic illness [43]. Some narratives also aligned with theoretical accounts of shame and blame in other partly-invisible conditions such as epilepsy [44].

More generally, participants’ concerns reflect the well-described phenomenon of ‘hidden disability’, which requires individuals to undergo a contextual negotiation about when to ‘pass’ as able-bodied, and when to self-identify as having a disability. In so doing they must weigh up conflicting drivers of self-identity and preservation of self, impression management, stigma, and legitimization of or possible value judgements based on illness-related behaviour [45, 46]. Moreover, the relapsing-remitting time course of brain fog symptoms also align with ‘episodic disability’, developed by people living with HIV to describe their experiences of unpredictable periods of wellness and illness [47], which adds an additional element of uncertainty to patients’ continual assessment.

Such requirements emphasize the extensive work which people with long Covid need to do to manage their condition and navigate services, which accord with theories of illness burden [33]. In particular, the communication and cognitive impairments compound the challenge of self-advocacy and system navigation in a healthcare system that has until recently lacked a clearly defined care pathway [6]. Accounts of positive experiences of care described established dimensions of good professional practice: active listening and bearing witness [40, 48]; wise counsel [39] and continuity of the therapeutic relationship [41] that alleviate patients’ illness burden and help begin to construct a healing narrative.

Lack of mechanistic understanding of the pathophysiological cause was a frequent frustration for participants. Ongoing research has hypothesized that neuronal damage during the initial illness secondary to direct viral neurotoxicity [49] or associated neuroinflammation generate a multisystem dysfunction resulting from a loss of central control and generalized peripheral inflammatory response [50]. Such suggestions are supported by pathological evidence of SARS-CoV-2 neurotropism [51] and neuroinflammation [52] combined with animal models of SARS-CoV-2 infection leading to neuroinflammation, intracellular Lewy body formation, or neuronal loss [25, 53]. It has been hypothesized that such processes impacting on vulnerable brain regions could correlate with neurocognitive long Covid symptoms: dysfunction of the brain stem, which is involved in regulation of both respiration and arousal – and thus potentially ‘brain fog’ -could result in the attentional deficits and disproportionate breathlessness seen in long Covid [54, 55], though this may not be the only explanation for the symptoms described in our empirical data.

Finally, our findings illustrate that whatever the explanation at the molecular and physiological level, the resultant impacts result from – and contribute to – a far wider interplay of psychological, physical and social factors. The clear disruption to an individual’s professional self, interpersonal relationships, and overall sense of identity, combined with the impact of a hidden and episodic disability impair sufferers’ abilities to achieve a previously anticipated state of ‘health’, described by Tarlov as ‘the capacity, relative to potential and aspirations, for living fully in the social environment’ [56]. Given that long Covid seems to be more prevalent amongst individuals of working age or those still in education, or amongst particular occupational ‘key worker’ groups who were at greatest exposure risk from Covid-19, the potential impact on society is highly significant. Therefore, whilst further work must deepen and exploit our mechanistic understanding, commissioners and providers of long Covid services, as well as individual clinicians, must remain cognizant of the disruption to these broader components of health and wellbeing and consider how they may best be mitigated.

### Strengths and limitations of the study

To our knowledge, this is the largest and most in-depth qualitative study of neurocognitive symptoms of long Covid published in the academic literature to date. The research team included both clinicians and social scientists. Our participants spanned a wide range of ages, ethnic and social backgrounds, and illness experiences – including, importantly, the under-researched majority who were never hospitalised. The majority of our participants became infected in the first wave of the pandemic, meaning they are among the earliest cohort of patients to experience long Covid, with email follow-up almost 12 months post-infection giving an insight into the natural history of the condition. We oversampled men and people from non-White ethnic groups to partially correct an initially skewed sample. The use of multiple linked sociological theories allowed to produce a rich theorisation of the lived experience of the illness and draw on that theorisation to produce principles and practical proposals for improving services. We included experts by experience (people with

The study does have some limitations. The entirely UK-based sample included a high proportion of people recruited from a support group for those with neurological symptoms (hence, likely to be more severely affected), and was not fully corrected for some demographic skews. In particular we may not have fully captured the perspectives of some minority ethnic groups or diversity in occupational classes. By pragmatically recruiting largely from social media, we may have introduced an element of selection bias. In the time since the first pandemic wave, knowledge and treatment of both acute and long Covid have altered substantially with medical research and patient advocacy (although with geographic variation, and thus inequality, in provision of and access to long Covid services in the UK), which may influence the experience of long Covid for people infected at later time points. It is likely that despite striving do democratic collaborative research *with* patients, we may not have fully grasped the lived experience or represented all voices.

### Comparison with previous empirical studies

Our findings of persistent, debilitating neurocognitive symptoms in people with long Covid are in alignment with several retrospective cohort studies [18] and online patient surveys [4, 8, 57, 58]. Our study adds further context to explore the functional and psychosocial impact of such symptoms, their interaction with physical symptoms, and mitigating efforts by patients.

Comparisons have been made between long Covid and other syndromes with neurocognitive dysfunction. Infection with SARS-CoV-1 [59], Epstein-Barr Virus, Coxiella burnetii, Ross River virus [60], and Borrelia burgdoferi [61] can result in similar impairments to concentration and memory, typically correlated with persistent fatigue. However, the challenge of unpicking the underlying aetiology of such symptoms is illustrated by the example of chronic fatigue syndrome/myalgic encephalomyelitis (CFS/ME), where difficulties with executive function, short-term memory, attention and word-finding are incorporated in the diagnostic criteria of both the UK National Institutes for Clinical Excellence [62], US Centers for Disease Control and Prevention [63], and International Consensus Group [64], but where the underlying cause remains unclear.

Moreover, examples such as HIV-associated neurocognitive dysfunction, which afflicts over 40% of people with chronic HIV infection [65], impairing learning, memory, attention, and executive function, suggests possible overlap across multiple chronic viral infections. A recent study in Nature illustrates how such higher order disruptions may be mediated on a molecular level through viral-associated perturbations in general cellular functions such as cortical excitatory synaptic signalling, choroid plexus disruption enabling peripheral T cell infiltration, and promotion of pathological microglial and astrocyte subpopulations [66]. All of these mechanisms – and others – will require further elucidation.

Both the partially hidden nature of the neurological disabilities experienced by long Covid patients and the extensive work required to manage these and navigate services may exacerbate the impact of the epidemiological distribution of persistent symptoms. Recent data from the Office for National Statistics demonstrated that self-reported long Covid was greatest in people aged 35-69 years, women, people living in the most deprived areas, those in health and social care occupations, and those with another activity-limiting health condition or disability [2]. As for the acute infection, long-term sequelae of Covid-19 infection are strongly impacted by socioeconomic determinants such as poverty and structural inequalities such as racism and discrimination [67], which may affect health beliefs, health-seeking behaviours, or the response of health services. Whilst not directly reported by participants in this study, further work to explore the impact of such determinants on long Covid epidemiology and interactions with health services will be crucial to mitigate the impact of associated disability.

### Conclusion: implications for services and further research

In dealing with Covid-19, it is crucial that health policy begins to shift from an acute disaster response to managing a chronic crisis. This study has brought neuroscientists together with qualitative researchers to try to align the subjective illness experience as directly described by patients with the objective disease models that underpin therapeutic options for ongoing ‘brain fog’ experienced by long Covid patients. The profoundly disabling, persistent impacts in some people revealed here adds weight to arguments that we need to prevent Covid-19 in order to reduce the long-term burden of this disease on patients, the health service, and the wider economy. Moreover, it is crucial to mitigate the impact for those already affected through a better understanding of the pathophysiological mechanisms of this neurotrophic virus and further exploration of the best approaches to support cognitive, psychological, and occupational restoration.

The strong positive and negative emotional touchpoints [38] described by individuals when their accounts are—respectively—believed or dismissed underscores the importance of the clinical relationship in which the patient is listened to, believed, and supported — particularly in primary care, which is likely to be the patient’s first point of contact [68, 69]. Furthermore, the varied nature of the severe impacts of brain fog identified in this study highlight the importance of ensuring that specialist services for this condition are accessible, easily navigable, comprehensive and interdisciplinary—for example incorporating (where necessary) assessment and rehabilitation from clinical psychologists and occupational therapists [7]. Our findings affirm those of a previous study (with a partially overlapping sample) to co-design quality indicators for long Covid services, which emphasised the importance of continuity, clinical responsibility, multidisciplinary input, patient involvement, and use of evidence-based guidelines [6].

## Data Availability

The data from this study is not openly available on a repository but we would be happy to discuss sharing with potential collaborators given the appropriate information governance.

## Acknowledgements

We thank the 50 participants for their interest and contributions, and two experts by experience for helpful comments on a draft of the paper: Sharon Taylor, child psychiatrist and honorary senior lecturer at the Central and North-West London NHS Foundation Trust and Imperial College School of Medicine and Clare Rayner, Independent Occupational Physician, Manchester. Alex Rushforth and Sietse Wieringa undertook interviews for the original study of long Covid.

## Contributors and sources

EL and TG conceptualised and designed the study. EL, LH, CC, and KP conducted focus groups. EL and CC led data analysis, with input from LH, TG and KP and produced a first draft of the results section. EL and CC wrote the first draft of the paper which was refined by all authors. LH provided research assistant support and conducted some interviews. ST and CR provided expertise by experience and knowledge of patient-led research. CC presented findings to long Covid patient participants with assistance from EL and TG. All authors contributed to refinement of the paper provided additional references. EL is corresponding author and guarantor. EL affirms that the manuscript is an honest, accurate, and transparent account of the study being reported; that no important aspects of the study have been omitted; and that any discrepancies from the study as planned (and, if relevant, registered) have been explained.

## Copyright

The Corresponding Author has the right to grant on behalf of all authors and does grant on behalf of all authors, a worldwide licence to the Publishers and its licensees in perpetuity, in all forms, formats and media (whether known now or created in the future), to i) publish, reproduce, distribute, display and store the Contribution, ii) translate the Contribution into other languages, create adaptations, reprints, include within collections and create summaries, extracts and/or, abstracts of the Contribution, iii) create any other derivative work(s) based on the Contribution, iv) to exploit all subsidiary rights in the Contribution, v) the inclusion of electronic links from the Contribution to third party material where-ever it may be located; and vi) licence any third party to do any or all of the above.

### Box 1.

**Definitions**

#### Planning

the mental process allowing individuals to choose necessary actions to reach a goal, ascertain the required order, assign tasks to cognitive resources, and establish a plan of action.

#### Decision making

the cognitive process resulting in the selection of a belief or a course of action from multiple possible alternative options,

#### Flexibility

the mental ability to adjust activity and content of the cognitive system i.e. enabling a switch between different task rules and corresponding behavioural responses, maintaining multiple concepts simultaneously and shifting internal attention between them.

#### Complex attention

a person’s ability to maintain information in their mind for a short time and to manipulate that information eg: to perform mental arithmetic calculations.

#### Selective sustained attention

the ability to focus on an activity or stimulus over a long period of time even if there are other distracting stimuli present.

#### Divided attention

the ability to attend to multiple different stimuli at the same time, thus responding to more than one demand from the surroundings i.e. enabling multi-tasking.

#### Processing speed

the time it takes a person to do a mental task i.e. the at which a person can understand and react to the information they receive from sensory inputs and generate a reaction.

#### Working memory

a cognitive system with a limited capacity, capable of temporarily holding information to enable reasoning and guiding decision-making and behaviour.

#### Procedural memory

a type of <u>implicit memory</u> that aids the performance of particular types of tasks without <u>conscious</u> awareness of previous <u>experiences</u> eg: stored motor programmes of particular well-rehearsed actions.

#### Autobiographical memory

a memory system formed from episodes recollected from an individual’s life that combines <u>episodic</u> (personal experiences and specific objects, people and events experienced at particular time and place) and <u>semantic</u> (general knowledge and facts about the world) memory.

#### Free recall

a common memory task requiring individuals to recall any items from a previously memorized list either immediately or following a delay.

#### Cued recall

As above, individuals are required to recall items from a previously memorized list but may be given cues (often semantic) to encourage this.

